# Does total triage and remote-by-default consulting impact vulnerable groups: A pilot study

**DOI:** 10.1101/2020.11.04.20220046

**Authors:** Aaminah Verity, Dharmendra Naidu, Victoria Tzortziou-Brown

## Abstract

**Background:** COVID-19 mandated a rapid and dramatic transformation of general practice. ‘Total Triage’ (TT), where all consultations should be triaged first, and ‘Remote-by-Default’ (RbD) consulting, where a clinician should consult remotely unless there is a “clinical exception”, were advised. It is unclear how these new ways of working were implemented in practice, and how they impacted vulnerable patients.

**Aim:** To assess the impact of TT and RbD on vulnerable patients and identify mitigation strategies.

**Design and Setting:** A mixed methods service evaluation in Lewisham, London, an area of high deprivation.

**Method:** Three parallel datasets were collected and analysed: Semi-structured interviews with stakeholders working with vulnerable groups and qualitative data from forums with black and ethnic minority patients, a survey of General Practitioners exploring implementation of TT and RbD, and a mystery shopper exercise reviewing access and messaging of ten practices.

**Results:** Barriers to access for vulnerable patients included challenges navigating the new model, difficulty engaging with remote consultations and digital exclusion. There was wide variation in messaging regarding changes to services and the practical application of TT and RbD. Potential solutions included clearer practice guidance and patient messaging, more consistent implementation, and identification and recording of patient access needs to enable better tailoring of care provision.

**Conclusion:** This pilot study identified perceived and actual barriers to accessing general practice for vulnerable patients following the rapid introduction of TT and RbD consulting. It proposes immediate steps to mitigate some of these impacts and highlights the need for further research in this area.

**How This Fits In:** The rapid and widespread adoption, recommended by NHS England, of total triage and remote-by-default consulting in general practice has yet to be evaluated. We provide a first look at how these changes are impacting those with historic difficulties in accessing primary care under the traditional GP model. We also provide some local recommendations that can be implemented easily at practice level and beyond, to mitigate the impact of these changes whilst making recommendations for further research to corroborate these findings widely.

## Introduction

The COVID-19 pandemic mandated rapid and widescale changes to general practice. To facilitate infection control during the lock-down period, NHS England mandated ‘Total Triage’ (TT), where all consultations required some form of triage, and ‘Remote-by-Default’ (RbD) consulting, where a clinician should consult remotely (by phone or video consultation) unless there is a “clinical exception” ^1^.

General practice teams rose admirably to the challenge of implementing this new model rapidly alongside providing outreach care to vulnerable and shielded patients. As this model remains in operation, it is unclear how TT and RbD has affected vulnerable patient groups and their access to care. There have been concerns around the quality of remote consultations, with a documented reduction in rapport and information gathering ^2^. In addition, there has been no centralised inequalities impact assessment of the above policies to date. At the same time, NHS England has asked local primary care teams to increase the scale and pace of work to reduce health inequalities, as part of the phase three COVID-19 recovery plan ^3^. More recently, NHS England wrote to GP teams and asked them to ensure that they were maintaining provision for face-to-face consultations ^4^.

Prior to COVID-19, vulnerable patient groups were more likely to have poor access to primary care and poorer health outcomes ^5,6^. COVID-19 has highlighted concerns about service inequalities within the NHS, with the most deprived in society both at highest risk of catching and dying from the disease and at highest risk of adverse health outcomes secondary to lockdown^7^.

Groups that are likely to be disproportionately affected at this time include those who already have difficulties in accessing care under a traditional model. These include: those experiencing homelessness, vulnerable migrants (refugees, asylum seekers and undocumented migrants), sex workers, gypsies and travellers, those recently leaving prison, those with addictions, those on low income with poor access to IT infrastructure, those with mental health problems, those with learning difficulties impacting social functioning, and victims of domestic violence among others.

This pilot study aimed to explore the impact of TT and RbD within general practice on vulnerable patient groups locally in Lewisham.

## Methods

Three parallel sets of data were collected in the London borough of Lewisham:

1. Interviews with stakeholders who work with vulnerable groups, and qualitative data from feedback forums with patients from black, Asian and minority ethnic (BAME) communities.
2. A survey of Lewisham GPs exploring the implementation of TT and RbD, and health care professionals’ perception of access.
3. A mystery shopper exercise reviewing access and messaging of 10 practices in the North Lewisham Primary Care Network (NLPCN).

### Qualitative Data

Qualitative semi-structured in-depth interviews were performed with stakeholders who worked directly with vulnerable service users. They were approached and consented to perform remote interviews by phone or using online meeting software. Recordings were made using a smartphone and data was transcribed using Otter.ai app and manually checked and corrected. Consent was taken for direct quotes and responses were anonymised. The interviewee list (Table 1) represented groups who work with vulnerable migrants, people experiencing homelessness and drug addiction, health inclusion services, and other support services for vulnerable groups. Interview questions can be found in Supplementary Appendix A.

**Table 1-.**
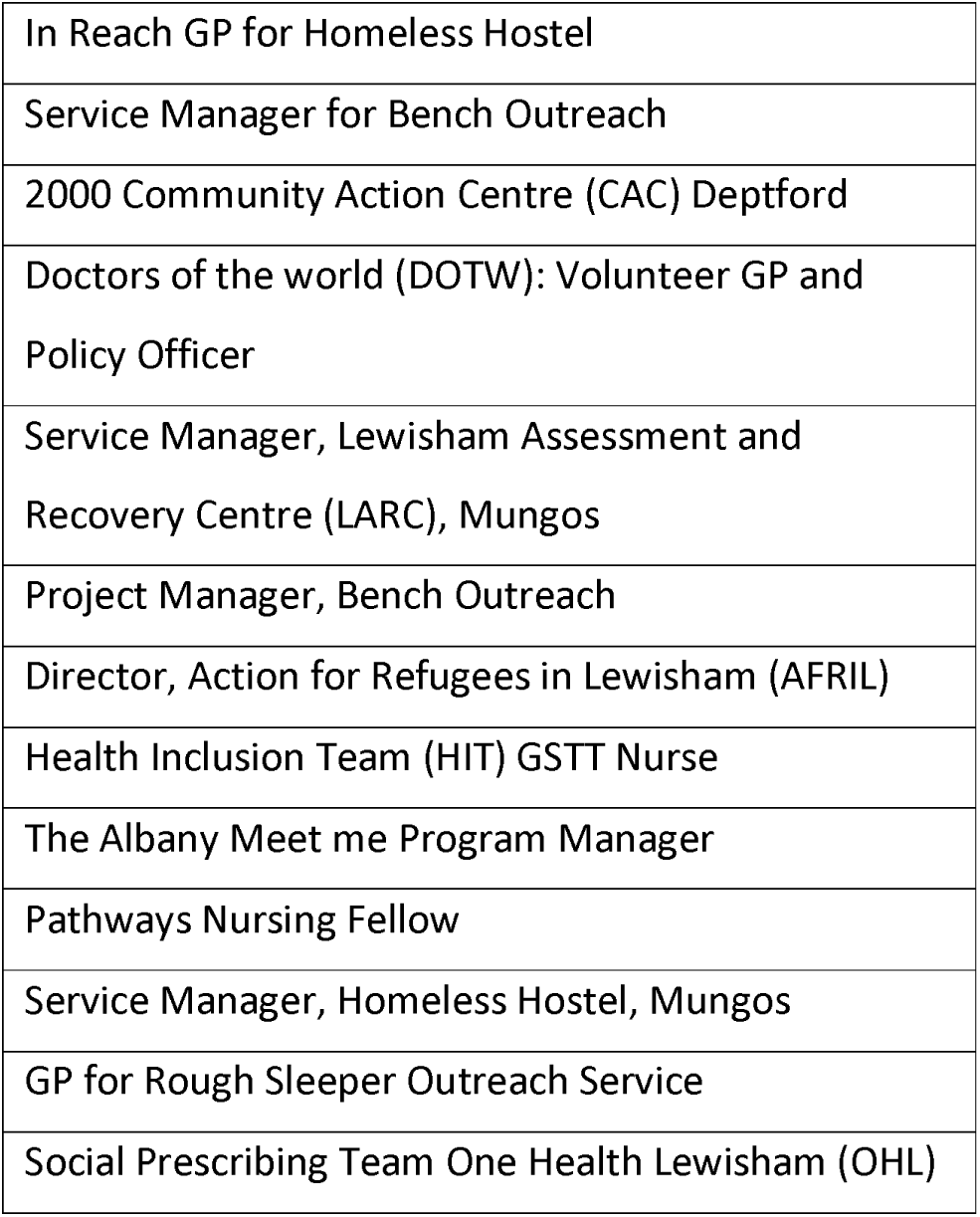
List of Interviewees for Qualitative Research

|  |
| --- |
| In Reach GP for Homeless Hostel |
| Service Manager for Bench Outreach |
| 2000 Community Action Centre (CAC) Deptford |
| Doctors of the world (DOTW): Volunteer GP and Policy Officer |
| Service Manager, Lewisham Assessment and Recovery Centre (LARC), Mungos |
| Project Manager, Bench Outreach |
| Director, Action for Refugees in Lewisham (AFRIL) |
| Health Inclusion Team (HIT) GSTT Nurse |
| The Albany Meet me Program Manager |
| Pathways Nursing Fellow |
| Service Manager, Homeless Hostel, Mungos |
| GP for Rough Sleeper Outreach Service |
| Social Prescribing Team One Health Lewisham (OHL) |

Transcriptions were reviewed and checked, and an inductive thematic analysis based on Braun and Clarke^8^was performed using QSR Nvivo version 1.2 for Mac. 21 Codes were created which were organised into 5 basic themes and then 4 overarching themes.

Healthwatch Lewisham were performing focus group discussions known as “Feedback Forums” aiming to understand the experiences of healthcare of BAME communities in Lewisham during the COVID-19 pandemic. Four forums were held with 21 participants from Lewisham. Participants were invited through advertisements by the Africa Advocacy Group, an organisation working with vulnerable groups. Two questions were added by the study author to the questionnaires used in the forums to assess the impact of TT. Results were transcribed, analysed and summarised by the Healthwatch Lewisham team and shared.

### Survey to GP Practices

The survey aimed to examine how practices in Lewisham have adopted TT and RbD consulting. The survey was piloted with GPs outside Lewisham to check that questions were comprehensible and that timing to complete survey was under 10 minutes. The survey was designed using Google forms and publicised through professional WhatsApp groups and through NLPCN mailing lists. The questions comprising the GP survey are shown in Supplementary Appendix B. The survey responses were analysed using Microsoft Excel and Google Forms.

### Mystery Shopper Exercise

A review of access and messaging for the 10 practices that make up NLPCN was performed following permission from the NLPCN clinical director. All practice websites were visited and practice phone numbers were called on the same day, and patient-facing messaging on service changes were documented. On a separate day, all practices were visited, entry to the practice was attempted and messaging on the door was recorded. Finally, all practices were called and asked to register a new patient without online access or forms of ID. Responses were recorded, summarised and anonymised.

## Results

### Qualitative Data

The interviews identified 4 overarching themes: Positive Experiences of TT, Barriers to Accessing GP services, Challenges of Remote Consulting, and Proposed solutions.

#### Positive experiences of TT

Several positive benefits of RbD and TT were identified. Greater appointment availability, ease of prioritisation of those needing urgent care and simpler interactions for those working as advocates for vulnerable patients. This shows the potential for this model to improve access to general practice as long as due consideration is given to overcoming the barriers it creates. For some groups who traditionally feel stigmatised in mainstream spaces, it was felt that remote consulting reduced some barriers to care.

> *“I think TT does present opportunity. Particularly engaging groups that find the GP practice environment itself intimidating, [swapping] for a place where they’re comfortable then, of course it can work”*

One respondent felt that there was more flexibility within the TT and RbD system which would work better with individuals with more chaotic lifestyles.

> *“So, before total triage you had to call in at 8am in the morning to try and get an appointment. So a lot of our clients were sort of frozen out, because that eight o’clock in the morning is geared towards somebody that’s got their life in some sort of order”*

#### Barriers to accessing GP services

The analysis showed that there were specific concerns surrounding how TT would entrench existing barriers to access and create new ones (Figure 1). Intrinsic barriers were identified including fears of mainstream services due to concerns around data confidentiality or worry about eligibility for services and the inability to prioritise health needs.

**Figure 1-.**
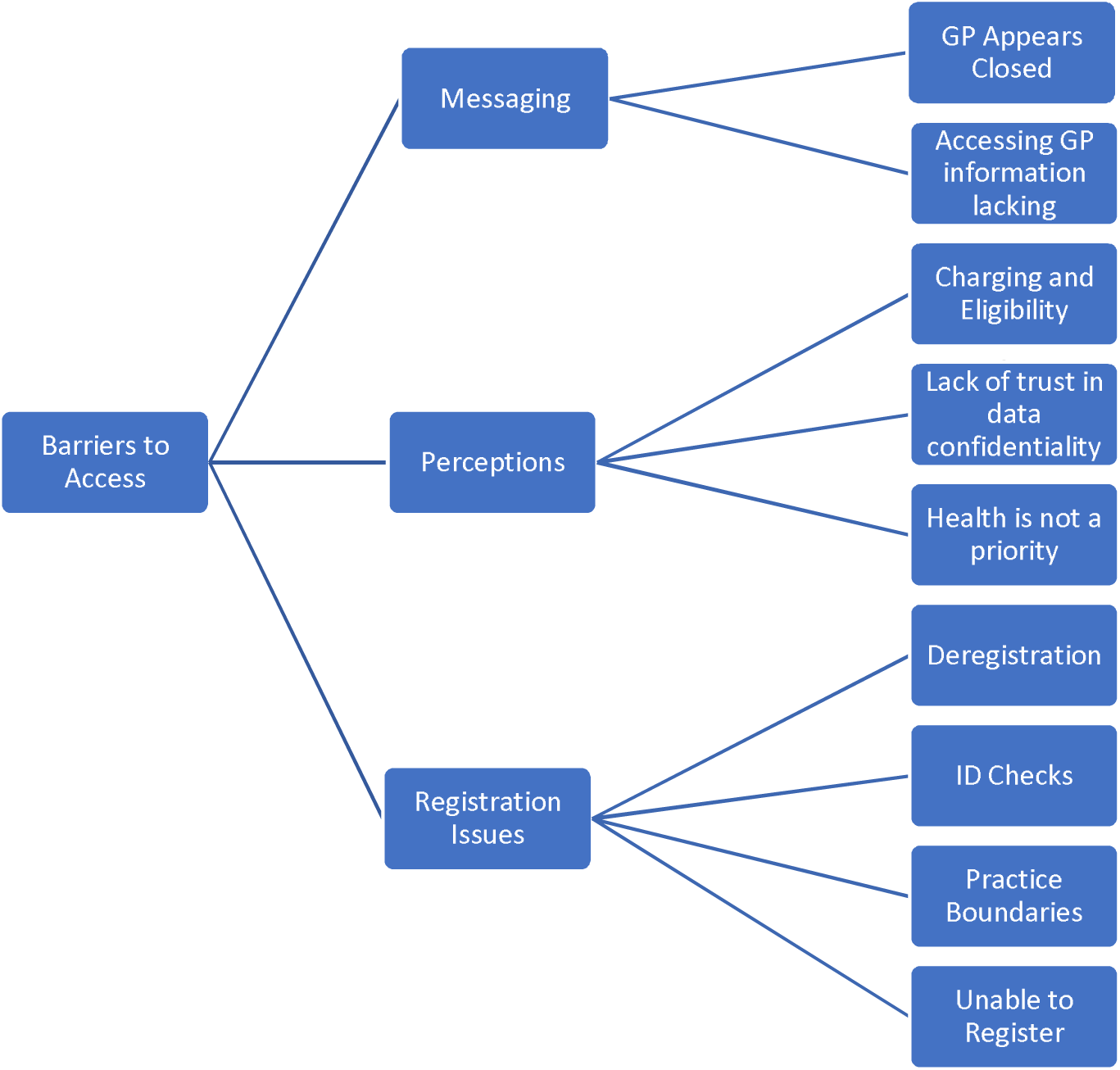
Barriers to Access thematic network

> *“I think people may be more scared about accessing services just because they get really uncomfortable giving certain details. I think doing that over the phone is more difficult because they feel like they can’t have a conversation”*
>
> *“They don’t prioritise their health needs. They don’t prioritise a lot of things that a regular person might do. I think that the TT system is difficult enough for a regular person to get through at times. So, I think for our clients it’s going to be a really, really big ask”*

There were specific concerns about new barriers to access through inconsistent messaging around the changes. The removal of the ability to walk-in to some practices has raised concerns amongst some service providers that those without advocates could be left out of services due to difficulties navigating the new system.

Regarding changes to the registration process under the new model, many stakeholders were initially optimistic that online registration may remove some barriers but in fact have found registration processes more laborious and feared that those without advocates would have no means to navigate the online systems and the subsequent ID checks being requested.

> *“Now, in COVID, you just look at that GMS 1 form, and it doesn’t ask for those things [proof of ID] and you think, I can just press submit and then that patient will be registered, but it’s seldom the case. We’re [then] being asked to think of ways that the patient could bring in their ID or can they scan it, or can we drop it through the door*.*”*

A new problem emerged around possible de-registrations because of lack of response when practices tried to contact vulnerable groups of patients at the beginning of the pandemic. The “Everyone In” campaign, to house the street homeless, involved some patients moving across London borough boundaries causing difficulties in maintaining registration with their previous practices.

> *“We’ve had a number of people deregistered because apparently, they were contacted in April and didn’t respond, and therefore they de-registered them. I said this person has been moved, because of COVID-19, because they’re homeless and now he needs the service”*

#### Challenges of Remote Consulting

There were three main themes (Figure 2) identified that prevented vulnerable groups from engaging with healthcare under a RbD model: having chaotic or harmful lifestyles where prioritising health was not possible, language barriers and a difficulty building rapport and trust via remote consulting. Illustrative quotes are presented in Table 2. 7/13 stakeholders felt that remote consultations would rarely work with the unpredictability of their service users’ day-to-day lives. This was mentioned more among those who ran services for those with drug addiction and homelessness, but not exclusively. Stakeholders highlighted that even with advocates they had witnessed very few successful remote consultations during the pandemic.

**Table 2-.** Interviewee comments on the challenges of remote consulting

|  |
| --- |
| *"In terms of remote consultations, the answer is that it went basically pretty badly. The list of people that would be interested in seeing him [the GP] would be a lot shorter than when he used to come and do face to face... and then the proportion of people he would successfully get through to on the phone, sometimes it was zero. The total number of remote consultations over the course of the two and a half months probably was only about 5, 6 or 7 successfully."* |
| *"We know that a lot of practices don't use an interpreter on registration... Without English as a first language, but because they're in front of them, there's things you can do with body language and can communicate to a degree that obviously is completely lost when you're not in front of somebody..."* |
| *"It's issues around trust, anxiety. A very considerable number of our residents have mental health issues. How easy is it to understand somebody [remotely]... doctors aren't good at explaining things to people anyway and the amount of additional frustration that can creep in if there's an accent on one end or the other, technical language, lines a bit dodgy...A lot of people are just going to say forget this, I'm just going to ignore my health problem..."* |
| *"I think when you're seeing someone with a broken finger, there's always more around that. And you can't get that from a telephone call."* |
| *"So, again, it's just making sure people have phones or being aware that their phone numbers change all the time. I mean I've worked with the service for seven years. I've got one client and she had 17 number changes in that seven years..."* |
| *"we had stories of people who would run out of credit just in the queue on hold to the GP. And if you're somebody that's seeking asylum for instance and you've got 35 pounds a week to live on, and access to phone credit and data might be a secondary priority to access to food and transport...if they have to spend half of their money for the week on credit and then it all disappears while they sit on hold."* |

**Figure 2–.**
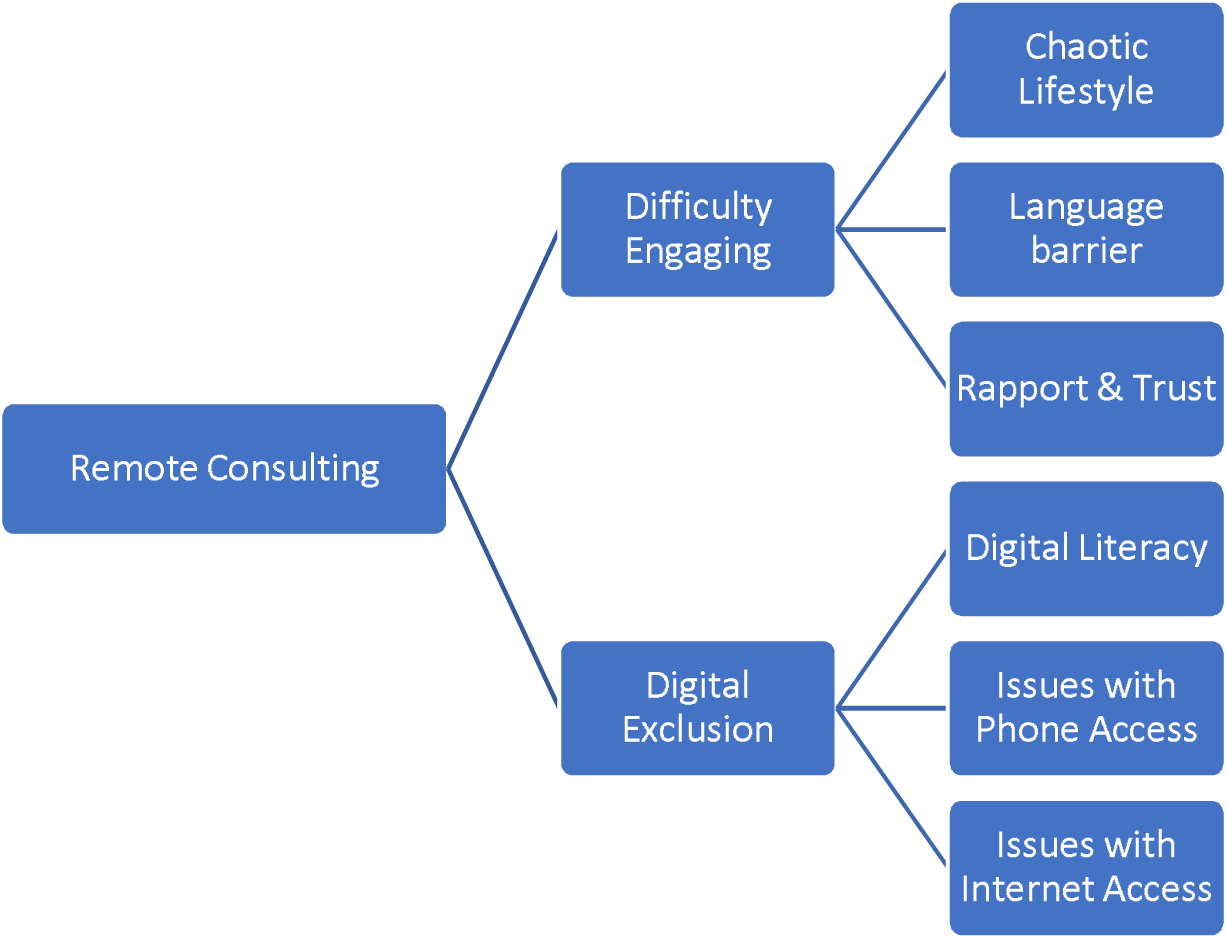
Issues with remote consulting thematic network

There was also concern from the Healthwatch forums around how triage models were being implemented. Patients felt uncomfortable giving clinical information to non-clinical staff and were also concerned about the ability of non-clinical staff to appropriately triage medical conditions.

Respondents (8/13) expressed concern that RbD consulting in general practice would negatively impact those with limited English where it was not seen to be common practice for GP receptionists to use translators even when clinicians did use them.

Respondents (7/13) felt that rapport and trust was not as easily attained through remote consultations. In addition, data from the Healthwatch Forums highlighted a reluctance for patients to share personal and medical details with non-clinical staff and worry about the confidentiality of non-clinical triage.

Digital exclusion was a significant concern highlighted by 10/13 respondents. Two types of digital exclusion were identified: a) those who lacked digital literacy or the ability to navigate remote consulting and b) those who through poverty were excluded through inability to maintain phone credit, data packages and IT infrastructure.

#### Potential solutions

When discussing potential solutions to overcome the barriers, five key themes were identified: clearer messaging on access options, identification of patient access needs during triage, accessing the practice by walking-in, patient advocates/translation services, and maintaining outreach services.

Clearer messaging on service changes and confidentiality, targeted at vulnerable groups through trusted sources, was seen as a way to overcome some of the barriers in place due to TT and RbD consulting.

Under a TT model several respondents suggested it would be important to ensure that when patients made initial contact with GP practices, call handlers made sure to ask how best to contact them and highlight access issues and preferences in patient records.

10/13 respondents mentioned that vulnerable patients must have the option to access the practice by walking-in if needed. It was acknowledged this should be done in a COVID-19 safe way as the pandemic continues.

Ongoing provision of face-to-face in-reach and outreach services for high need, vulnerable patients for whom mainstream services are not suitable, facilitation of the use of patient advocates and closer working relationships with them were seen as essential.

> *“We also made a recommendation to primary care to prioritize face-to-face appointments for those who face digital exclusion*.*”*
>
> *“The in-reach service has been invaluable. That direct access to a GP with almost zero notice, with admin taken away and often the resident isn’t directly involved in the conversation…without that we would actually be very concerned for the health and wellbeing of our residents*.*”*

### GP Survey Findings

There were 27 unique responses which represented 18/37 GP practices in Lewisham. GPs who responded were more weighted towards North Lewisham, but all four neighbourhoods were represented.

The majority (77%) of practices in Lewisham surveyed have moved to a TT system at some point during the COVID-19 pandemic. Most had clinician or reception-based triage models with 2/27 stating solely digital triage (Figure 3).

**Figure 3-.**
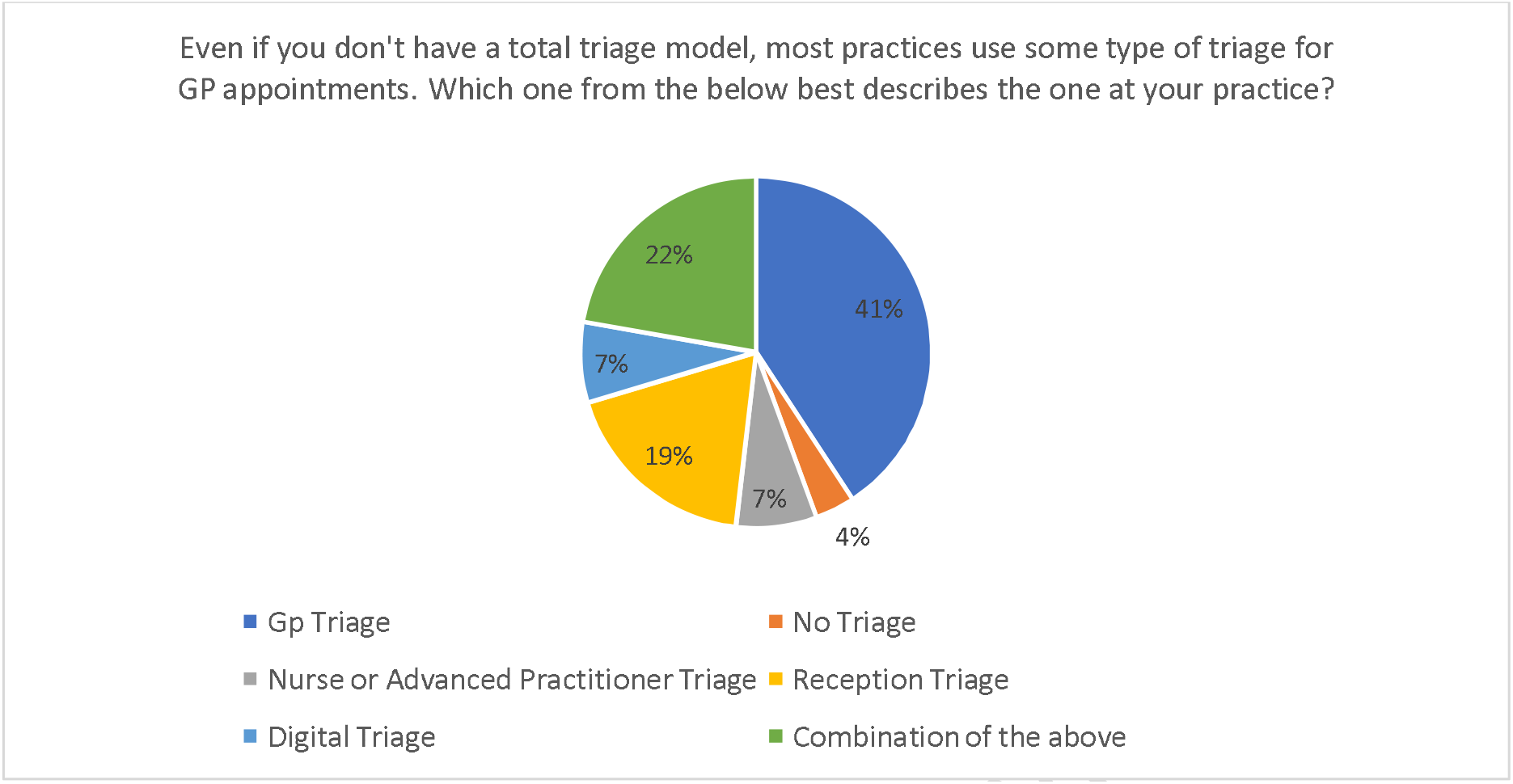
Survey results for question: “Most practices use some type of triage for GP appointments. Which one from the below best describes the one at your practice?”

Most respondents (60%) felt that triage was conflated with consultation, meaning clinicians were expected to deal with the problem there and then, rather than making another appointment with an appropriate healthcare professional. The majority (62%) of respondents had on average between 16-20 patient contacts in a clinical session, 19% had 21-25 contacts, and 15% had up to 15 contacts. One respondent stated they had 31-35 patient contacts per session.

Just over half (55%) of respondents had a method of online consulting, although in most cases the number of consultations that were purely online was less than 20%. 40% of respondents said that their practices had online-only patient registration.

With regards to continuity of care, 40% of respondents felt that TT had reduced this, while nearly 20% felt continuity had improved. Most respondents (85%) stated they had the ability to tailor the length of consultation for patients with complex needs and similarly 80% had a system in place to contact those who were difficult to reach remotely, including text messaging and letter writing.

The majority of respondents (22/27) expressed concern about reduced access for vulnerable groups under TT and RbD. 7/27 suggested maintaining walk-in provision for those who need it with a system of alerting staff by assessing patients’ ability to access services and flagging any issues using alerts on notes.

Finally, the survey showed significant variation in the implementation of TT, online and remote consulting in a small number of similarly geographically located practices. For example, 2/27 used digital only triage while 5/27 used reception only triage and 15/27 used online consultations while the rest had no digital consulting.

### Mystery Shopper findings

Full results of the mystery shopper exercise can be found in Supplementary Table 1. The findings showed that all practices advertised that they were open to new patient registrations: all had a method of online booking and 7/10 had an option for online consulting, whilst maintaining normal phone access.

However, there were some findings that corroborated concerns raised in the qualitative data. In general, phone messages were long and complex when attempting to explain the change in how to access care. Three messages lasted more than 2 minutes before the caller entered a call waiting system. Only 2/10 practices had a direct alert on their website explaining that since COVID-19 there had been changes to the booking system. There was some disparity within practices between the messaging on the website, the practice door and the phone message, providing a potentially confusing picture.

Regarding registration, ID checks were requested by 3/10 practices online and 4/10 of the practices could not assist with the registration of a patient with no internet access or ID. In 5/10 practices, receptionists facilitated registration by asking the patient to attend face-to-face when remote options were not feasible.

The majority of practices (6/10) were fully accessible to patients, but 4/10 did not allow walk-in patients and although there was messaging on how to access the practice, this was in English only. Bells and intercoms were available in 3/4 practices which did not allow walk-in patients. Signage stating “STOP do not enter” was prominently displayed in 7/10 practices, which, although necessary to alert patients to recent COVID-19 infection control measures, could be misunderstood by patients with vulnerabilities and/or limited English.

As with the GP survey, the mystery shopper exercise highlighted the significant variation in the implementation of the new policies in a small number of similarly geographically located practices.

## Discussion

### Summary

COVID-19 has shone a light on existing health inequalities and has mandated many changes to provision of healthcare ^7^. One adaptation that is likely to persist post-Covid-19 is the widespread adoption of TT and RbD consulting in general practice, originally introduced as a means of reducing exposure to the virus along with a governmental commitment to this model ^9^. GPs rose admirably to the immense challenge of shifting to a new model of care and implemented rapid changes to prevent GP surgery waiting rooms becoming hotbeds of transmission.

The findings of this study show that service users, GPs in Lewisham and those who provide support services for vulnerable groups have concerns around the impact of RbD and TT on access to general practice. The mystery shopper exercise and the survey substantiated several of the concerns identified from qualitative research. Specifically, concerns around difficulties registering, digital exclusion, barriers to remote consulting and inconsistency of messaging regarding changes to services since COVID-19 have been highlighted.

### Strengths and Limitations

This study provides three data sets triangulating experiences of GPs on the frontline with experiences of those that provide support services to vulnerable groups and an objective review of practices. Study limitations include the scope of this small-scale pilot study. The findings are not generalisable until large scale research is performed to include other vulnerable groups, conducted over a larger geographical area and including the voices of experts by experience.

### Comparison with Existing Literature

This study echoed findings from the DOTW Rapid Needs Assessment^10^, the Groundswell Listen Up paper^11^and the Patients Not Passports Briefing Paper “Migrant Access to Healthcare During the Coronavirus Crisis”^12^suggesting vulnerable groups have struggled to access healthcare during the pandemic.

There are benefits of the new consulting model especially when there is a need for prioritising access to care. Respondents echoed findings that, when done well, triage can be a useful tool to allocate resources appropriately and according to clinical need. Patient queries that can be dealt with by allied health professionals can free up GP time to deal with patients with complex needs^16^. E-consults have been associated with high levels of patient satisfaction, especially around response times, but their impact on quality of care and workload for GPs has yet to be fully elucidated ^17 18^.

This study highlights the importance of considering how best to implement triage systems without increasing inequalities in access. Concerns from patient groups around who is performing clinical triage is an important consideration. Patients’ concerns about confidentiality and sharing personal information with non-clinical staff were found to be exacerbated by this consulting method. These fears may become even more of a barrier to access with vulnerable migrants who may have worries about data sharing and hence choose not to consult if asked personal questions at triage, especially by non-clinical staff ^15^.

In the absence of central direction, a wide amount of variation was identified in approach to TT and RbD consulting, even within a small geographic region linked through a primary care network. There is a need to fully characterise this variance and elucidate why certain practices have adopted different implementations of TT and RbD. 76.9% of practices studied moved to TT during the pandemic. There are examples of practices who moved to this model prior to the pandemic, who adopted safeguarding procedures for vulnerable patients ^13^. Only recently, patient-facing messaging templates have been made available by NHS England to highlight changes to services ^14^. Practices need to be given adequate support and guidance in order to be able to provide an equitable and safe system for all patients.

In addition to concerns over inequality of access, this study also unveiled some concerns about the nature of remote consulting. There is evidence that remote consulting achieves poorer outcomes, producing more consultations, less information sharing and less rapport building ^2 19 20^. In addition, telephone triage has been associated with an increase in the number of primary care contacts which may explain the reported increase in sessional workload in some practices of our study ^21^.

E-consulting and remote consulting require different skills and can promote an “episode of care approach” rather than a holistic assessment of a patient and relational care. 40% of respondents to our GP survey felt that the triage model they were using had led to reduced continuity. Patients with complex medical needs, poor insight into their health conditions and those with vulnerabilities are reliant on an open-ended consultation, and proponents of E-consulting state that it is not appropriate for these patients ^22^. Although TT and RbD does not preclude vulnerable patients from making face-to-face appointments, there is a need for measures to ensure that the new GP systems do not present obstacles for those who cannot prioritise or advocate for their own health needs.

### Implications for Research/Future practice

This pilot study examines the potential impact of TT and RbD consulting in general practice for vulnerable service users and identifies initial steps which can be taken to address perceived and actual barriers to access at a local level. The potential actions that can start to mitigate the concerns highlighted by this study are summarised in Box 1. Adequate funding and central guidance will be needed for some of these. Some of the actions may also apply in secondary care settings where remote consulting has also been widely and rapidly adopted.

Rapid commissioning of research exploring the impact of these new ways of working on access for vulnerable groups of patients and ways of addressing any negative consequences is urgently needed. In the meantime, existing recommendations within the DOTW Safe Surgeries Toolkit^23^, Migrant’s Access to Healthcare During the Coronavirus Crisis^12^, the Listen Up Report on Digital primary care by Groundswell^11^and the Pathway GP receptionist standards ^24^could mitigate some of these effects and the recommendations from this paper are being adopted by the primary care network in North Lewisham.

#### Box 1

Recommendations for improving access for vulnerable patients under TT and remote by default consulting

- ***Clear and consistent messaging*** for practices and for patients across all mediums explaining how to access services under RbD and TT, dispelling myths on closed practices and lack of face-to-face appointments, informing about registration and reassuring about confidentiality.
- ***Reducing the length of time on call waiting***, or ***provision of a freephone number or call back service*** when contacting practices.
- ***Access to interpreters*** both at reception and for consultations.
- Providing a ***triage system which considers patient’s disparities in access***, flagging these for future interactions and making adjustments according to patient needs.
- ***Promoting continuity of care, face-to-face appointments where needed and adjusting appointment length for vulnerable groups of patients with complex needs***.
- ***Working closely with patient advocates*** to facilitate access to primary care for those who cannot advocate for themselves.
- Maintaining an ***outreach and in-reach primary care service*** for those groups who are unable to engage with mainstream services.

## Data Availability

The data that support the findings of this study (suitably anonymised) are available on request from the corresponding author.

## Ethical Approval

QMUL Research Governance Team have advised the project is classified as non-research as it is a local study/service evaluation and therefore ethical approval is not required.

## Competing interests

The authors have no competing interests to declare. A declaration of our other interests can be found at:

http://www.whopaysthisdoctor.org/doctor/432/active

http://www.sunshineuk.org/doctor/888/active

## Acknowledgements

The authors would like to thank the North Lewisham Primary Care Network for their support of this project.

We would also like to thank Pathway, especially Samantha Dorney-Smith, Nursing Fellow for their support.

Our sincere thanks to Healthwatch Lewisham who agreed to collaborate to add questions to their feedback forums.

We would also like to thank those who contributed their time to our interviews, GP survey and the practices who participated in the mystery shopper.

