## Appendices for "Does total triage and remote-by-default consulting impact vulnerable groups: A pilot study"

### Appendix A: Interview Questions for Stakeholders

#### Interview for Stakeholders

***Does Total Triage (TT) in primary care disproportionately impact those from vulnerable groups?***

This interview has been designed to identify the impact of the TT model of primary care on access to care for vulnerable patients in South East London. Your answers are anonymous but the organisation you represent will be quoted. We will be recording and transcribing the interview, so we can analyse what was talked about.

As a key stakeholder working with vulnerable groups we are interested in hearing your assessment of how the move to TT in primary care has affected your client's access to primary healthcare.

Please try to answer as fully as possible. We are especially interested in any innovative ideas you may have for improving access for vulnerable patients at this time.

For further information or any questions about the project please contact

**Interviews for focus groups/informants**

- TT
  - What do you understand by the TT model of primary care?
  - How do you think this has impacted your clients?
- Access to care
  - Have you heard any reports of problems accessing primary care since surgeries moved to this model?
  - Can you think of any specific examples where TT has provided an insurmountable obstacle?
  - What type of care is being impacted most? E.g. urgent primary care, medication requests, admin support like sick notes
  - Which elements of primary care are actively reaching out to your clients?
- Continuity of care
  - Have clients been able to continue contacting their GP for routine management of long term conditions?
  - Have any clients faced interruption to medication supply?
- Registration
  - Are you aware of any clients who have been unable to register since we've moved to TT?
  - Are you aware of any practices that are deregistering patients who are temporarily out of borough due to lockdown?
- Digital exclusion
  - How is your organisation overcoming issues with patients who don't have phones or internet access?
- Proposed solutions
  - Any examples of practices where care is still going well?
  - What model of primary care would work for your clients in a post covid world?
  - Do you think clients need walk in centres without triage?

Preprint Under review

### Appendix B: Questions for GP Survey

#### Impact of Total Triage on access to Primary Care for Vulnerable patients

##### Exploring the impact of new triage systems post covid on provision of care for vulnerable patients

*Required

1. Practice Name *
2. Job Role * Mark only one .

    Gp Partner, Salaried Gp, Practice Manager, Gp Registrar, Practice Nurse, Admin Staff

3. Are you currently operating under a total triage model i.e. No patient can make an appointment without first being triaged? *

    Yes, No

4. When did you implement your total triage system?

    Prior to Covid-19 pandemic, During the pandemic, Have not moved to total triage model, Other:

5. Even if you don`t have a total triage model, most practices use some type of triage for GP appointments. Which one from the below best describes the one at your practice? *

    Reception triage (receptionist books a phone appointment or face to face GP appointment after triaging)

    GP triage (GP triages the patient and advises on need for further appointments)

    Digital triage first (Patients are asked to complete an online triage form to access a clinician)

    Nurse or Advanced Practitioner triage (Non Doctor clinician triages patients and advises on need for further appointments)

    No triage (patients can book only a phone appointment without being triaged e.g. online booking)

    Combination of the above

    Other:

6. If your practice uses GP triage, does it differentiate between triage (i.e. quick assessment of the problem so that the patient is given the right consultation appointment) and consultation (i.e. full assessment and management of the problem with shared decision making as appropriately)

    Yes, we have separate triage and consultation slots. When triaging clinicians are not expected to deal with the problem there and then unless this is a very quick administrative task.

    No, triage is conflated with consultation and clinicians are expected to deal with the problem there and then most of the time rather than making another appointment.

    Not relevant as my practice does not have GP triage

    Other:

7. What is the maximum number of patient contacts in a clinical session (4 hours and 10 minutes) that a salaried doctor is expected to have? (This includes all patient contacts e.g. face to face, telephone, e-consultations, video).

    1-15, 16-20, 21-25, 26-30, 31-35, 36-40, 41-50, >50, Other:

8. Are you using a method of online consulting? *

    Yes, No

9. If yes, what proportion of your consultations are only online?

    <5%, 5-19%, 20-49%, 50-79%, >80%

Preprint Under review

### Provision of Care

10. How do you think the shift to more remote working (remote consultations) has affected relational continuity of care at your practice? *

Improved continuity, Reduced continuity, Has not influenced continuity, Other:

11. Do you have the flexibility to tailor the length of patient appointments to their needs (e.g. book a double appointment for patients with complex needs or if there is a need for an interpreter)?

Yes, for the vast majority of cases (>90%)
Yes, for 50-90% of cases
Only for 20-49% of cases
Rarely, for <10 % of cases

12. Does your practice have a system of using different methods of contacting patients who don't respond/miss appointments and need to be seen? (e.g. writing letters to those who do not answer their phones). *

Yes, No, Don't Know, Other:

13. If yes, what is the system?

14. How is your practice managing new patient registrations? Check all that apply

Online only, In person with form, Don't know, Other:

### Proposed Solutions

15. Do you have any ideas how access to primary care would look in Lewisham in a post covid world, for patients who cannot go online or have difficulty using phones?

Preprint Under review

Appendix C: Anonymised Mystery Shopper Data Set

| Practice | Website Message | Registration info on website | Message on door | Physical Entry | ID check | Phone message | Appt booking | Registration attempt |
|---|---|---|---|---|---|---|---|---|
| 1 | We are currently welcoming new patients | "You should come along to reception with your medical card and fill in the registration form. If you do not have a medical card, we can still accept you with a proof of address." | Surgery door is closed but surgery is open for consultation please call for any query "please do not come the surgery, telephone and discuss the reason" | Door Locked need to call to be let in | Yes | No phone message. | Book on phone, or Patient access AskNHS | Called – bring in ID to practice and fill out form. Stated no ID. Suggested go to another practice. |
| 2 | To book an appointment please telephone the surgery. | Please complete the below forms and submit to (email) | Our surgery doors are closed, if you have an appointment call us on the usual number, you will be asked to wait outside to be collected by the clinician you are seeing. If you do not have an appointment you can reach us on the usual number or download and register on our app | Door Open | Not mentioned | 2mins. Please do not visit the surgery if you have not spoken to a GP first | Book on phone, Ask NHSapp or patient access | Called – directed to website. No internet access told to come and buzz at the door say no internet and they can give physical form |
| 3 | X is still here for you, our lines are still open and you | Just complete the form below and we'll have you registered within 2 | STOP only patient with confirmed appointments may enter the surgery. | Door open | No | Only patients with confirmed pre-booked appointments are | Book by phone, online or email | Called and told to use website No internet told to come to |

Preprint Under review

| | | | | | | | | |
|---|---|---|---|---|---|---|---|---|
| | can still book appointments. | working days. It's simple, it's quick and once it's done, it's done. | Please sanitise hands, put on gloves, check temperature and check in | | | allowed to enter the surgery | App | practice knock on window and they will give form and register |
| 4 | No mention of changes to access or COVID-19 | Download form and send to practice or print off and hand in | STOP please do not enter the practice unless you have been invited by a doctor to do so. | Door Locked, ring doorbell for entry | On form states photo ID checked | No phone message | Book by phone or online, no mention not to come to practice | Called told to use website, pick up form and bring back with ID, told no ID said just fill out form and come back next day |
| 5 | No mention of not being able to register | Register online redirects to patient access | Stop please do not enter practice, if you need any assistance please knock on the door and somebody will assess you | Door Locked knock For entry | No form | COVID-19 information about travel history. Around 2 mins | Econsult, patient access, Phone | Called redirected to online. Told no internet told to try another practice. |
| 6 | We are still open just working differently SEL CCG video | Register online fill out these questions – asks you are ordinarily resident in the UK | To protect our patients and our staff face coverings must be worn if you have to attend the GP surgery | Open | Not mentioned | Still here to help you just in different ways, ask NHS app to book telephone or video appt, | Ask NHSapp, patient access, online, phone | Called redirect to online Told not accepting paper registrations and need to get help to go online |
| 7 | COVID-19 Important notice for all | Register online form on website, button on homepage | Coronavirus stay at home save lives, please do not enter this | Door locked, | Not mentioned | Telephone lines open and travel info for covid risk | Call after 8am, online | Redirected to online. Come to |

Preprint under review

| | | | | | | | | |
|---|---|---|---|---|---|---|---|---|
| | patients Please be advised, because of the current situation we have changed our face to appointments to telephone consultations with immediate effect. | | building unless you have been advised to. Any queries please ring the bell | entry by intercom | | | access using patient access | surgery and give paper form |
| 8 | COVID-19 page says stay at home if you have symptoms no mention on homepage of change to services | Online form<br><br>Please fill in your details and click Submit when complete. | Practice is open, However, to keep our patients and staff SAFE, we ask that you go online or call. Only enter the practice if you have been asked to do so | Open | No | Chest pain or severe breathlessness call 999, phone lines are busy,<br>You have reached X stay at home for 7 days if cough or fever etc, advise to go online to use econsult >2 mins | Call<br>Econsult<br>NHS App<br>Patient access | Redirected to online<br>Told no paper option because of COVID19 currently |
| 9 | No mention of change to services or COVID-19 | If you live in or near our catchment area, we'd love for you to join our practice. | STOP only patient with confirmed appointments may enter the surgery. | Door open | No | Only patients with confirmed pre-booked appointments can | Online patient access<br>Call | Redirected to online.<br>Come down and get a form. |

Preprint Under review

| | | | | | | | | |
|---|---|---|---|---|---|---|---|---|
| | | Just complete the form below and we'll have you registered within 2 working days. | Please sanitise hands, put on gloves, check temperature and check in | | | enter the surgery, all non-urgent care being provided by telephone and video consultations | Email<br><br>Online consult: no | |
| 10 | Covid banner redirect to NHS website for covid info | We are currently welcoming new patients. We operate an "open list" facility.<br>In order to register you must complete a Patient Registration Form, available at reception or download if you prefer | Due to coronavirus if you need a face to face appointment you will need to speak with a doctor first, please call to arrange a consultation for any other queries please speak to a receptionist | Open | Yes ID and address verification | If you wish to book a face to face appt you will need to speak to a doctor first when transferred please your name and contact information and you will be asked for a reason for your call | Patient access<br>Ask NHS app<br>Phone | Come to surgery and collect form and then bring back with ID – told no ID they want ID, said I have none told they will check with doctor |

Preprint Under review
